# Automated Interpretation of Fundus Fluorescein Angiography with Multi-Retinal Lesion Segmentation

**DOI:** 10.1101/2024.12.20.24319428

**Authors:** Ziwei Zhao, Shoujin Huang, Weiyi Zhang, Fan Song, Yan Lu, Xianwen Shang, Mingguang He, Danli Shi

## Abstract

**Purpose:** Fundus fluorescein angiography (FFA) is essential for diagnosing and managing retinal vascular diseases, while its evaluation is time-consuming and subject to inter-observer variability. We aim to develop a deep-learning-based model for accurate multi-lesion segmentation for these diseases.

**Methods:** A dataset comprising 428 standard 55° and 53 ultra-wide-field (UWF) FFA images was labeled for various lesions, including non-perfusion areas (NPA), microaneurysms (MA), neovascularization (NV) and laser spots. A U-net-based network was trained and validated (80%) to segment FFA lesions and then tested (20%), with performance assessed via Dice score and Intersection over Union (IoU).

**Results:** Our model achieved Dice scores for NPA, MA, NV, and Laser on 55° FFA images at 0.65±0.24, 0.70±0.13, 0.73±0.23 and 0.70±0.17, respectively. UWF results were slightly lower for NPA (0.48±0.21, p=0.02), MA (0.58±0.19, p=0.01), NV (0.50±0.34, p=0.14), but similar for Laser (0.74±0.03, p=0.90). Notably, NV segmentation in choroidal neovascularization achieved a high Dice score of 0.90±0.09, surpassing those in DR (0.68±0.22) and RVO (0.62±0.28), p<0.01. In RVO, NPA segmentation outperformed that in DR, scoring 0.77±0.25 versus 0.59±0.22, p<0.01, while in DR, MA segmentation was superior to that in RVO, with scores of 0.70±0.18 compared to 0.53±0.20, *p*<0.01. Moreover, NV segmentation was significantly stronger in venous phase (0.77±0.17) and late phase (0.75±0.24) compared to arteriovenous phase (0.50±0.32), p<0.05.

**Conclusion:** This study has established a model for precise multi-lesion segmentation in retinal vascular diseases using 55° and UWF FFA images. This multi-lesion segmentation model has the potential to expand databases, ease grader burden and standardize FFA image interpretation, thereby improving disease management. Furthermore, it enhances interpretable AI, fostering the development of sophisticated systems and promoting cross-modal image generation for medical applications.

**Synopsis:** We developed deep-learning models for segmenting multiple retinal lesions in both normal and ultra-field FFA images; the satisfactory performances set the foundation for quantifiable clinical biomarker assessment and building interpretable generative artificial intelligence.

## Introduction

Retinal vascular diseases, including diabetic retinopathy (DR), retinal vein occlusion (RVO) with its subtype branch retinal vein occlusion (BRVO), and neovascular age-related macular degeneration (nAMD) coupled with choroidal neovascularization (CNV), pose a substantial global health challenge. As leading causes of vision loss and blindness, these diseases affect millions and impose considerable personal and socioeconomic costs.^1^ Specifically, DR prevalence in diabetics is 27.0%, contributing to 0.4 million cases of blindness,^2^ and the projected incidence of AMD by 2040 is 288 million, with about 10% expected to develop neovascular complications.^3^

Fundus fluorescein angiography (FFA) is a critical diagnostic tool to identify retinal vascular abnormalities such as microaneurysms (MA), neovascularization (NV), and capillary non-perfusion areas (NPA). Segmenting and quantifying these lesions are critical for accurate diagnosis, disease staging, and monitoring. For instance, in DR, quantifying NPAs is crucial for deciding on interventions; larger NPAs indicate more severe ischemia and a higher risk of disease progression, potentially requiring photocoagulation treatment.^4^ MAs are significant in various retinal vascular diseases.^5^ In DR, they are crucial for early detection and risk assessment, where their dynamics predict disease trajectory. ^6–8^ Although previously underestimated in RVO, recent studies show that MAs, especially in collateral vessels, are key indicators of persistent retinal edema in BRVO, underscoring their value in these scenarios.^9,10–11^ MAs are most visible on FFA, and their quantification is useful to identify patients at higher risk of vision-threatening complications, necessitating closer monitoring and proactive treatment.^12^

However, accurate interpretation of FFA images requires ophthalmologists to have specialized knowledge and training. In remote areas, doctors may not possess the requisite expertise to produce reliable FFA reports, even though facilities are equipped for the examination.^13^ In urban settings, clinicians face time constrains, needing 5-10 minutes to analyze and report each FFA examination, increasing their workload.^14^ Even among trained professionals, there is a notable variation in interpreting FFA reports, which becomes more problematic in multi-center studies due to inconsistent manual annotations.^15^ Inconsistencies in FFA analysis can further impact the use of quantification tasks to guide interventions or assess prognosis.

The advancement of artificial intelligence (AI) has made it a valuable tool in medical diagnostics. Previous research has explored the application of deep learning (DL) techniques for DR grading and lesion detection.^16^ Pan et al.’s study achieved multi-label detection in DR using FFA, with sensitivities of 79.7% for NPA, 98.0% for MA, 84.0% for leakage, and 80.3% for laser scars.^17^ However, their focus has predominantly been on image-level lesion classification without specific localization or delineation. Deep learning technology including convolutional neural networks (CNNs) has shown promise in segmenting FFA lesions, as detailed in Table 1. These studies reveal DL’s potential for accurate lesion identification in FFA images of specific disease datasets.^14,15,18–24^ Nevertheless, limitations exist: research has mostly concentrated on FFA images of specific angles —— 30°, 55° or ultra-widefield (UWF), and targeted specific lesions within certain retinal diseases such as NPA in DR,^15,18,23^ NPA in BRVO,^19,21^ and CNV in nAMD.^24^ This narrows the scope of lesion detection across diverse diseases.

**Table 1:**
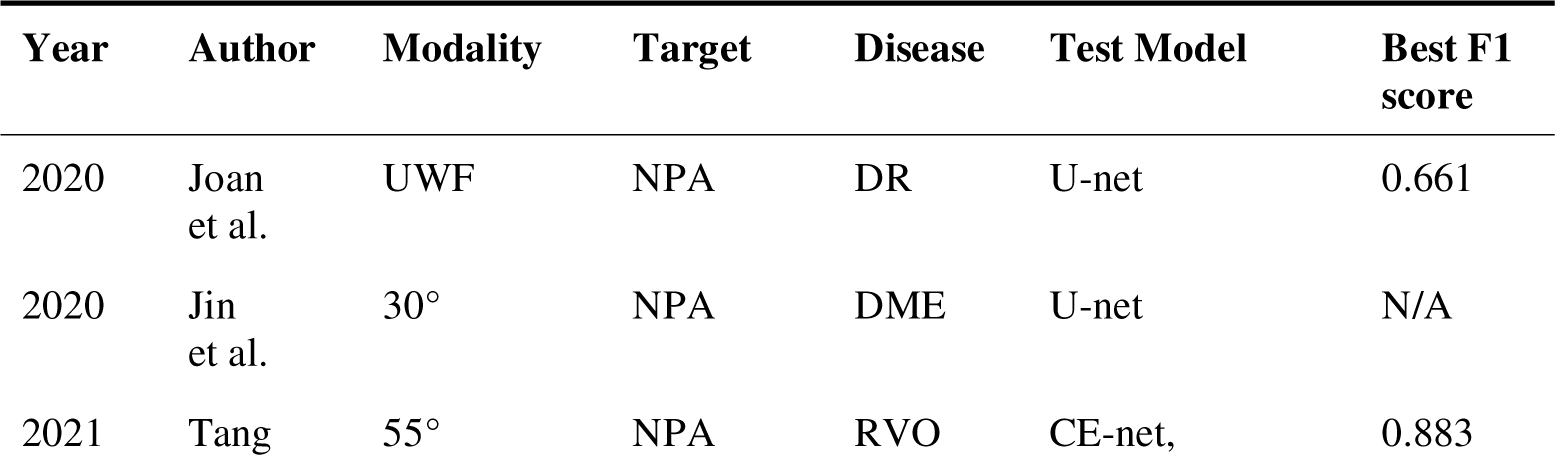
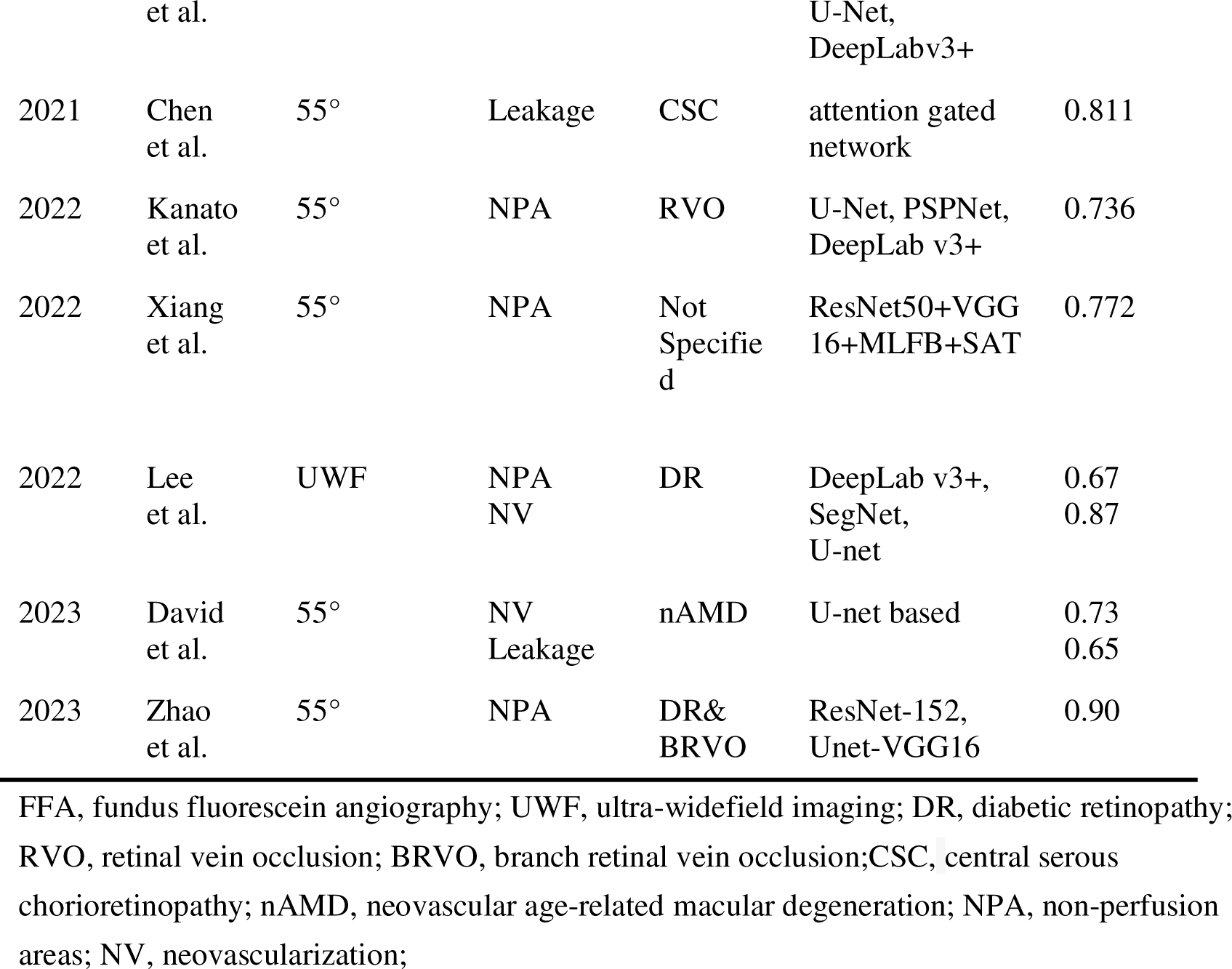
Summary of previous studies on lesion segmentation performance in FFA images.

To address the research gap, our study aims to tackle the challenges of manual interpretation and the necessity for lesion quantification in retinal vascular diseases. Initially, we have gathered a substantial repository of 55°and UWF FFA images, carefully annotated with NPA, MA, NV, and post-photocoagulation changes. Furthermore, we have trained an AI model that can identify these changes in various diseases. Overall, we propose an automated, objective methodology for detecting and segmenting common lesions in FFA images. This innovation aims to enhance the accuracy, efficiency, and consistency of retinal disease evaluations across a spectrum of retinal vascular conditions.

## Materials and Methods

The flow chart of the study is shown in Figure 1..

**Figure 1.**
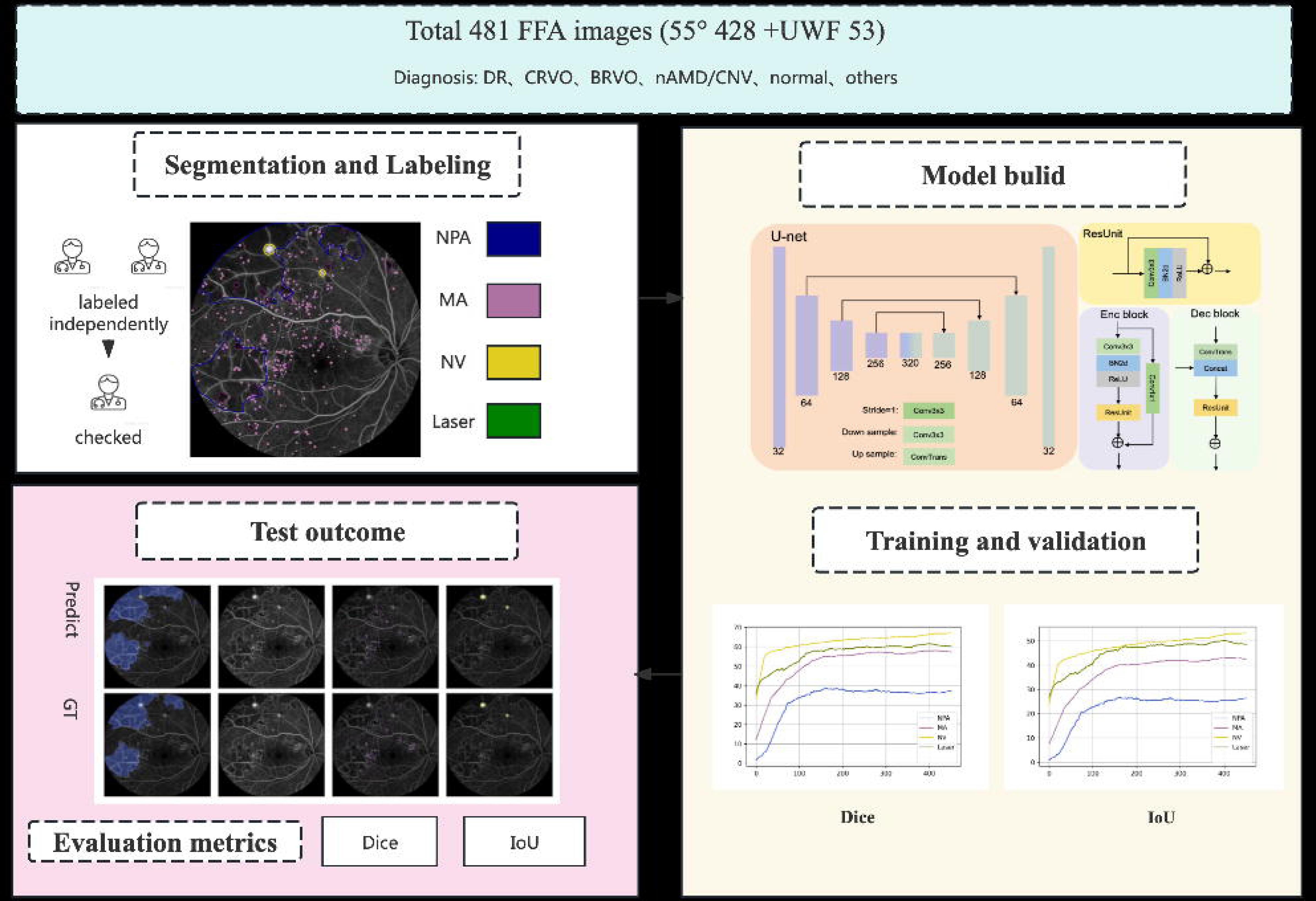
Flow diagram of this study.

### Dataset

The dataset was collected from multiple tertiary hospitals, comprising 428 55°and 53 UWF images. 55° images were captured using Zeiss FF450 Plus (Carl Zeiss Meditec, North America) and Heidelberg Spectralis (Heidelberg Engineering, Heidelberg, Germany) cameras at a resolution of 768 × 768 pixels. Additionally, to conduct a preliminary exploration of the model’s performance on UWF images, 53 UWF images were included. UWF images were captured by the Optos California (Optos plc, Dunfermline, Scotland, UK) at a higher resolution of 1536 × 1024 pixels. Our dataset features a spectrum of retinal conditions, including DR, CRVO, BRVO, nAMD, CNV, normal eye, and other diseases. It also covers various phases: arterial, arteriovenous, venous, and late. We excluded images from different phases of the same eye to prevent data overlap. Most images, including those with regionally obscured retinal areas due to hemorrhages, were retained to represent the diversity of clinical practice. Only images of extremely poor quality—where retinal structures were nearly indiscernible and lacked clinical reference value—were excluded. All patient data were anonymized and individual consent was waived. The Institutional Review Board of the Hong Kong Polytechnic University approved the study (HSEARS20240301004).

### Labeling and Image Preparation

Two experienced ophthalmologists (Z.Z. and F.S.) annotated the dataset using Labelme (*Ver 3.16.2,*) independently. For each image, graders utilized the polygon tool to outline the optic disc, macular region, non-perfusion areas (NPA), neovascularization (NV), and laser spots (Laser), while the point tool was applied to identify microaneurysms (MA). They also determined the most likely diagnosis and the phase. NPAs were defined as regions with an intercapillary distance exceeding normal levels but lacking fluorescein filling. NV appeared as highly fluorescent areas with an irregular branching pattern, sometimes accompanied by leakage. Following the methodology of Joan, non-informative peripheral areas in UWF images were excluded by defining a ‘gradable area’—a zone on UWF-FFA images where lesions and anatomical structures are discernible.^15^ The ground truth (GT) was established through consensus between the 2 graders and confirmed by a retinal specialist.

Before feeding the input image into the network, we performed Min-Max normalization to scale them within the range of 0 to 1. We implemented a robust data augmentation method, applying a series of techniques including Gaussian modifications, brightness and contrast adjustments, resolution reduction, and gamma shifts, each with assigned probabilities of 10%, 15%, 25%, and 10% respectively. Furthermore, random horizontal and vertical flips were applied to both the images and their corresponding labels. These data augmentation methods improved detail capture and model generalization.

### Network Architecture

Our network architecture is constructed based on the U-net framework,^25^ incorporating residual units^26^ in both the encoding and decoding stages as shown in Figure 1. For the encoding stage, input features undergo downsampling through 3×3 convolutions with a stride of 2. Subsequently, these features pass through N iterations of the residual structure, with the input features being summed with the final result. During the decoding stage, features from the skip connections are concatenated with the input features, following N iterations of residual structures. Ultimately, the decoder progressively restores the encoded features.

Note that instance normalization and ReLU activation functions are applied following all convolutional operations. The channel variations are as follows: 32, 64, 128, 256, 320. At each upsampling/downsampling stage, repetitions of residual units are three. The network is configured with 1 channel input and 10 channels output. For the segmentation task, we treat the five resulting segments as five binary segmentation tasks.

### Training and testing details

During the training phase, we employed the SGD optimizer with the Poly learning rate decay algorithm. The learning rate was set to 0.01, with a weight decay of 3e-5, batch size of 1, and momentum of 0.99. The u-net model was trained for 500 epochs, and the selection of the model was based on the best mean dice score of the validation set. The loss function utilized was a combination of dice loss and cross-entropy loss. The loss function is computed separately for each of the four categories, and the individual losses are then aggregated to obtain the final loss used for backward propagation. The model was trained on one GPU with Tesla V100 and CPU with Intel(R) Xeon(R) Gold 6248R CPU.

### Performance metrics

For image inference, we implemented a testing augmentation strategy, involving random horizontal and vertical flips applied to the input image into the network to obtain diverse outcomes, respectively. Subsequently, the predicted probability values were averaged to derive the final segmentation result.

The performance of the model was quantitatively evaluated using two common metrics for image segmentation tasks: the Dice score and the Intersection over Union (IoU).

1. The Dice score measures the similarity between the predicted and actual segmentations. It is defined as:

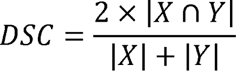 Here, X and Y represent the predicted and ground truth pixel sets, respectively. The Dice coefficient ranges from 0 (no overlap) to 1 (perfect agreement).
2. The IoU metric evaluates the overlap between the predicted segmentation and the ground truth. The formula is:

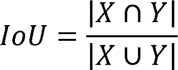

Similar as the Dice score, X and Y are the predicted and ground truth pixel sets. IoU values span from 0 to 1, with 1 indicating perfect overlap.

## Results

### Dataset Compilation

Our dataset comprised 428 55° FFA images and 53 UWF-FFA images, totaling 481 FFA images. For a focused study, we prioritized the 55° images due to their larger quantity, while also including UWF images for preliminary model performance evaluation on a smaller sample size. For the 55° FFA images, 305 were allocated for training, 35 for validation, and 85 for testing. Correspondingly, the UWF FFA images were divided into 40 for training, 3 for validation, and 10 for testing.

### Distribution of Diseases

The dataset comprised a diverse retinal pathologies: 286 images of DR, 19 of CRVO, 47 of BRVO, 48 of nAMD or CNV, 10 of normal fundus, and 20 of other conditions requiring further examination.

### Label Frequency

Label frequency analysis of the dataset indicated the presence of 1,527 ‘NPA’, 57,975 ‘MA’, 459 ‘NV’, and 12,762 ‘Laser’. The mean values for each lesion were 3.2 ‘NPA’, 120.5 ‘MA’, and 0.95 ‘NV’ labels. The pixel frequencies for ‘NPA’, ‘MA’, ‘NV’, and ‘Laser’ were 0.088, 0.006, 0.008, and 0.021, respectively. The frequency and pixel count of each label in 55° and UWF FFA images are detailed in Table 2.

**Table 2.** Distribution of label frequency and pixel counts in 55° and UWF FFA images.

| Label name | Frequency counts |  |  | Pixel counts |  |  |
| --- | --- | --- | --- | --- | --- | --- |
|  | 55° | UWF | All | 55° | UWF | All |
| NPA | 1024 | 503 | 1527 | 20946340 | 8731017 | 29677357 |
| MA | 41741 | 16234 | 57975 | 1519689 | 486512 | 2006201 |
| NV | 359 | 100 | 459 | 2305044 | 388791 | 2693835 |
| Laser | 9509 | 3253 | 12762 | 5659824 | 1257354 | 6917178 |

### Performance on 55°FFA Images

Model performance metrics, including the Dice and IoU scores for 55°, UWF FFA, and the overall dataset are presented in Table 3. The detection of NPA, MA, and NV on 55° FFA images achieved comparable levels to previous studies, with Dice scores of 0.65±0.24, 0.70±0.13, and 0.73±0.23, respectively. In targets not previously studied, namely Laser, the Dice score was 0.70±0.17.

**Table 3.**
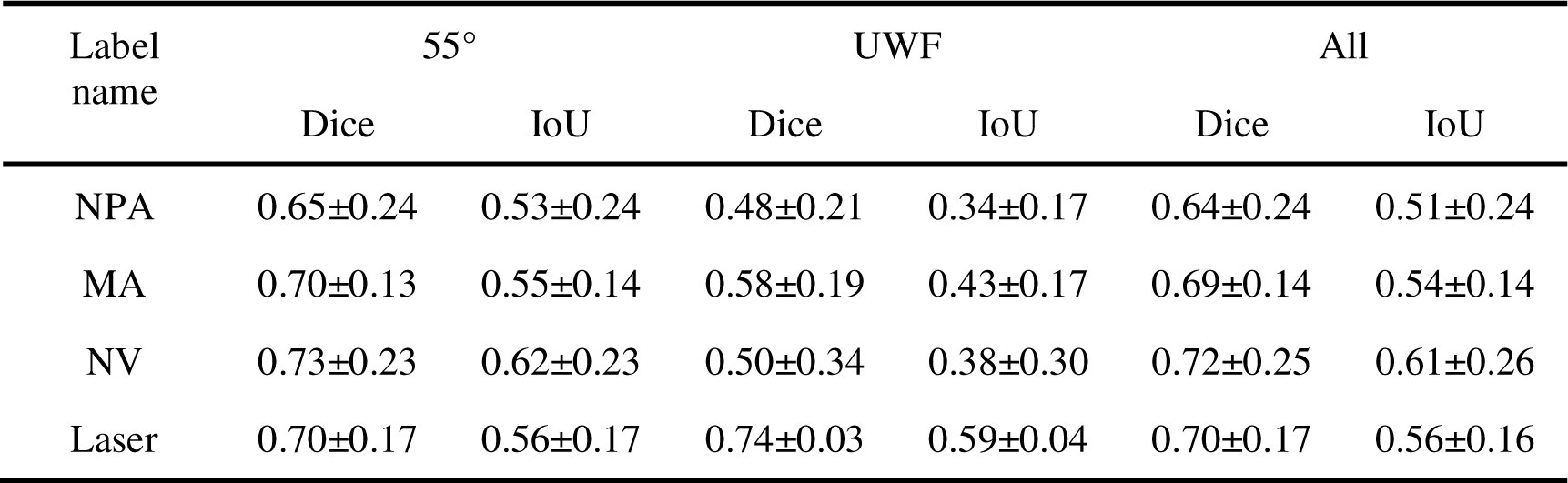
Model performance metrics on 55° FFA, UWF FFA, and the overall dataset.

| Label name | 55° |  | UWF |  | All |  |
| --- | --- | --- | --- | --- | --- | --- |
|  | Dice | IoU | Dice | IoU | Dice | IoU |
| NPA | $0.65 \pm 0.24$ | $0.53 \pm 0.24$ | $0.48 \pm 0.21$ | $0.34 \pm 0.17$ | $0.64 \pm 0.24$ | $0.51 \pm 0.24$ |
| MA | $0.70 \pm 0.13$ | $0.55 \pm 0.14$ | $0.58 \pm 0.19$ | $0.43 \pm 0.17$ | $0.69 \pm 0.14$ | $0.54 \pm 0.14$ |
| NV | $0.73 \pm 0.23$ | $0.62 \pm 0.23$ | $0.50 \pm 0.34$ | $0.38 \pm 0.30$ | $0.72 \pm 0.25$ | $0.61 \pm 0.26$ |
| Laser | $0.70 \pm 0.17$ | $0.56 \pm 0.17$ | $0.74 \pm 0.03$ | $0.59 \pm 0.04$ | $0.70 \pm 0.17$ | $0.56 \pm 0.16$ |

### Preliminary Glimpse on UWF-FFA Images

The segmentation performance of NPA, MA, and NV on the UWF dataset was lower than that on the 55° dataset, as seen in Table 3. The Dice scores for NPA, MA, and NV decreased slightly to 0.48±0.21(p=0.02), 0.58±0.19(p=0.01), and 0.50±0.34(p=0.14), respectively. The Dice score of Laser remained at a comparable level to the 55° dataset, recorded at 0.74±0.03(p=0.90).

### Results by Disease Type

In DR, the Dice scores for NPA, MA, NV, and Laser were 0.59±0.22, 0.73±0.09, 0.66±0.25, and 0.70±0.11, respectively. In contrast, for RVO, the corresponding Dice scores were 0.77±0.25, 0.53±0.20, 0.71±0.29, and 0.70±0.31. Additionally, in CNV, the Dice score for NV was notably high at 0.90±0.09. Table 4 details Dice and IoU scores for each label by disease. Figure 2 highlights the differences in Dice scores, demonstrating superior segmentation performance for NPA in RVO compared to in DR, and better MA segmentation in DR over in RVO (p<0.01). NV segmentation performed best in CNV, outperforming both DR (p=0.02) and RVO (p=0.22).

**Figure 2.**
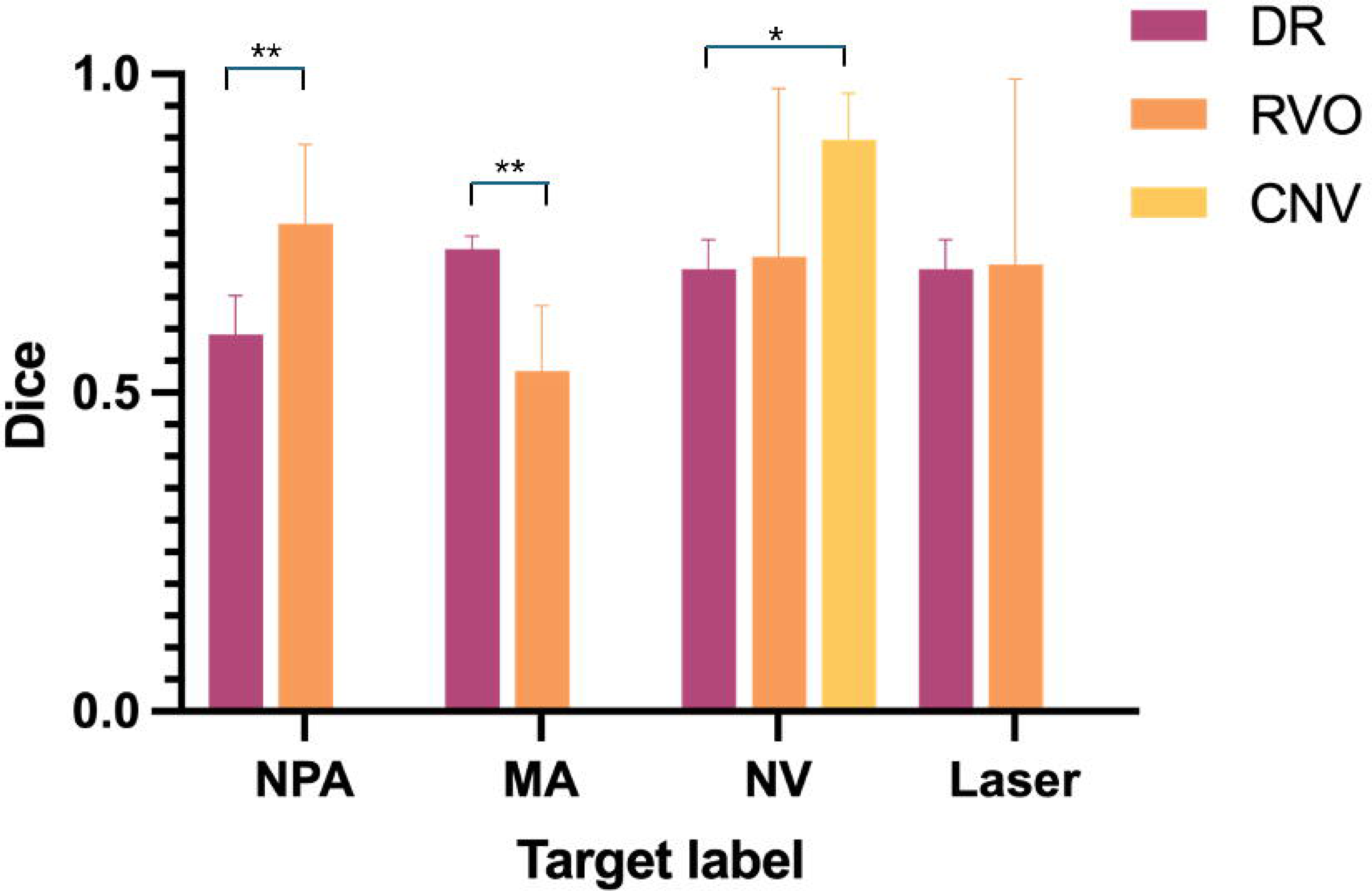
Comparative dice scores for target labels across DR, RVO, and CNV.

**Table 4.** Model performance metrics by disease category.

| Label<br>name | DR |  | RVO |  | CNV |  |
| --- | --- | --- | --- | --- | --- | --- |
|  | Dice | IoU | Dice | IoU | Dice | IoU |
| NPA | $0.59\pm0.22$ | $0.45\pm0.20$ | $0.77\pm0.25$ | $0.67\pm0.28$ | — | — |
| MA | $0.73\pm0.09$ | $0.55\pm0.14$ | $0.53\pm0.20$ | $0.43\pm0.17$ | — | — |
| NV | $0.66\pm0.25$ | $0.54\pm0.26$ | $0.71\pm0.29$ | $0.61\pm0.29$ | $0.90\pm0.09$ | $0.82\pm0.14$ |
| Laser | $0.70\pm0.11$ | $0.54\pm0.11$ | $0.70\pm0.31$ | $0.60\pm0.28$ | — | — |

### Results by Phase

As illustrated in Figure 3, segmentation of NPA, MA, and Laser showed no significant differences across the phases: A-V (arteriovenous), V (venous), and late. However, for NV, segmentation capability was significantly stronger in phases V (Dice=0.77±0.17, p=0.01) and late (Dice=0.75±0.24, p=0.02) compared to phase A-V (Dice=0.50±0.32). Detailed model performance metrics by phase, are provided in Table 5.

**Figure 3.**
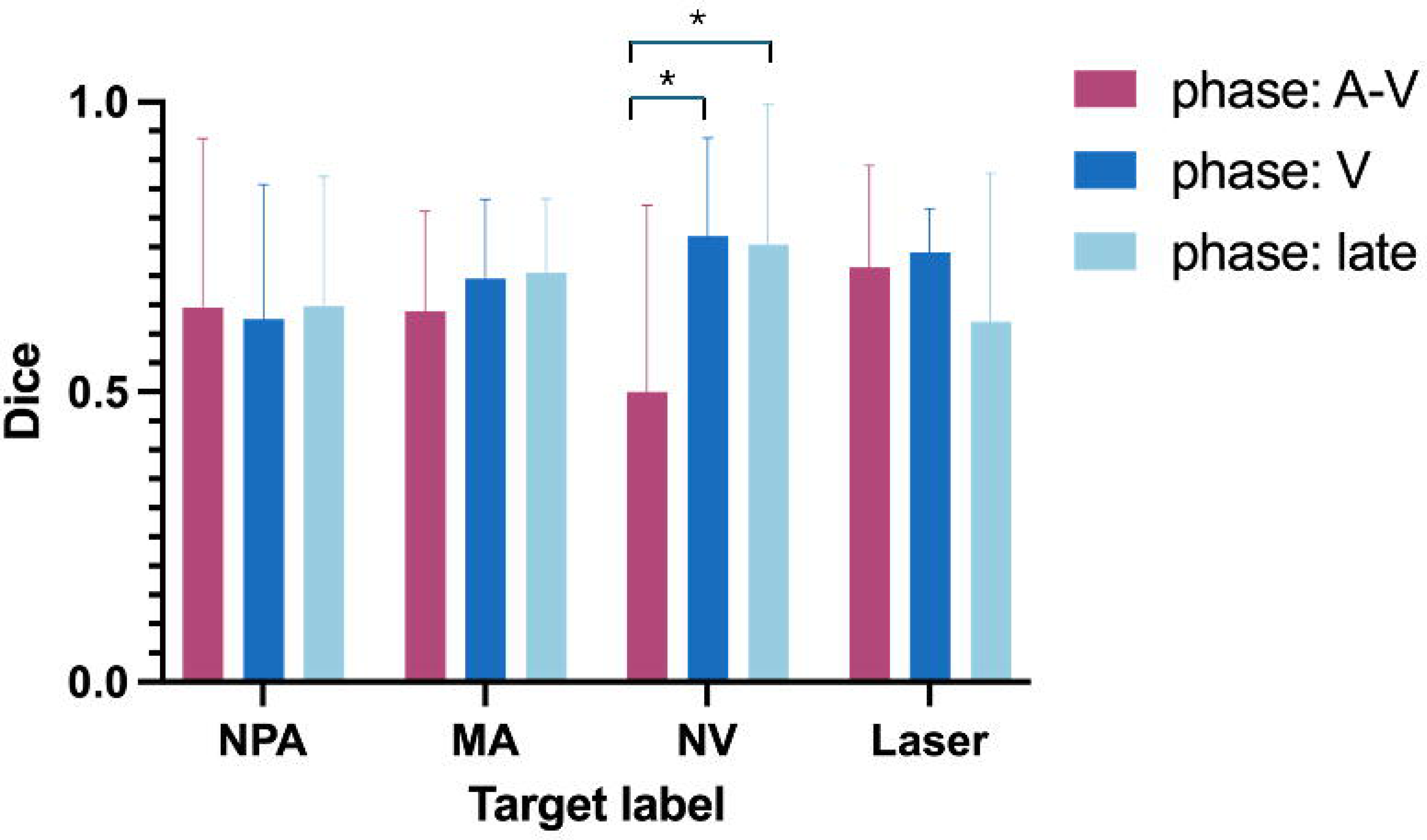
Comparative dice scores for target labels across arteriovenous, venous, and late phases.

**Table 5.** Model performance metrics by phase.

| Label<br>name | Phase: A-V |  | Phase: V |  | Phase: late |  |
| --- | --- | --- | --- | --- | --- | --- |
|  | Dice | IoU | Dice | IoU | Dice | IoU |
| NPA | 0.65±0.29 | 0.53±0.29 | 0.63±0.23 | 0.49±0.23 | 0.65±0.22 | 0.51±0.23 |
| MA | 0.64±0.17 | 0.49±0.16 | 0.70±0.14 | 0.55±0.14 | 0.71±0.13 | 0.56±0.14 |
| NV | 0.50±0.32 | 0.39±0.32 | 0.77±0.17 | 0.65±0.21 | 0.75±0.24 | 0.65±0.26 |
| Laser | 0.71±0.18 | 0.58±0.19 | 0.74±0.08 | 0.59±0.09 | 0.62±0.26 | 0.49±0.22 |

### Qualitative Evaluation

We present model predictions and ground truth visualizations for labels in 55° FFA images, shown in Figures S1–S5. Overall, the segmentation performance for different lesions is commendable. Recognition of NPA is particularly strong, especially in BRVO cases. MA predictions generally align well with GT, though there are false positives, such as confusing parts of laser spots or BRVO-related hyperfluorescence with MA. The model demonstrates high accuracy in identifying typical NV but falls short in the early stages or subtle leakage, occasionally mistaking strong vascular leakage as NV. Laser spot segmentation is generally satisfactory, despite lower Dice scores in atypical cases. Some predictions even identified laser spots missed in our GT. In UWF FFA images (Figure S6), the model exhibits less accurate NPA detection in peripheral areas, with MA and NV performance not on par with the 55° FFA images, although segmentation of laser spots remains consistent.

### Ablation study: Impact of Training Set Size on Model Performance

We further segmented the original training set into distinct subsets, each constituting 20%, 40%, 60%, and 80% of the full dataset, to evaluate the impact of training set size on model performance. The results, as depicted in Figure 4, illustrate that the average model scores for the labels NPA, MA, NV, and Laser consistently improved with lager training set sizes.

**Figure 4.**
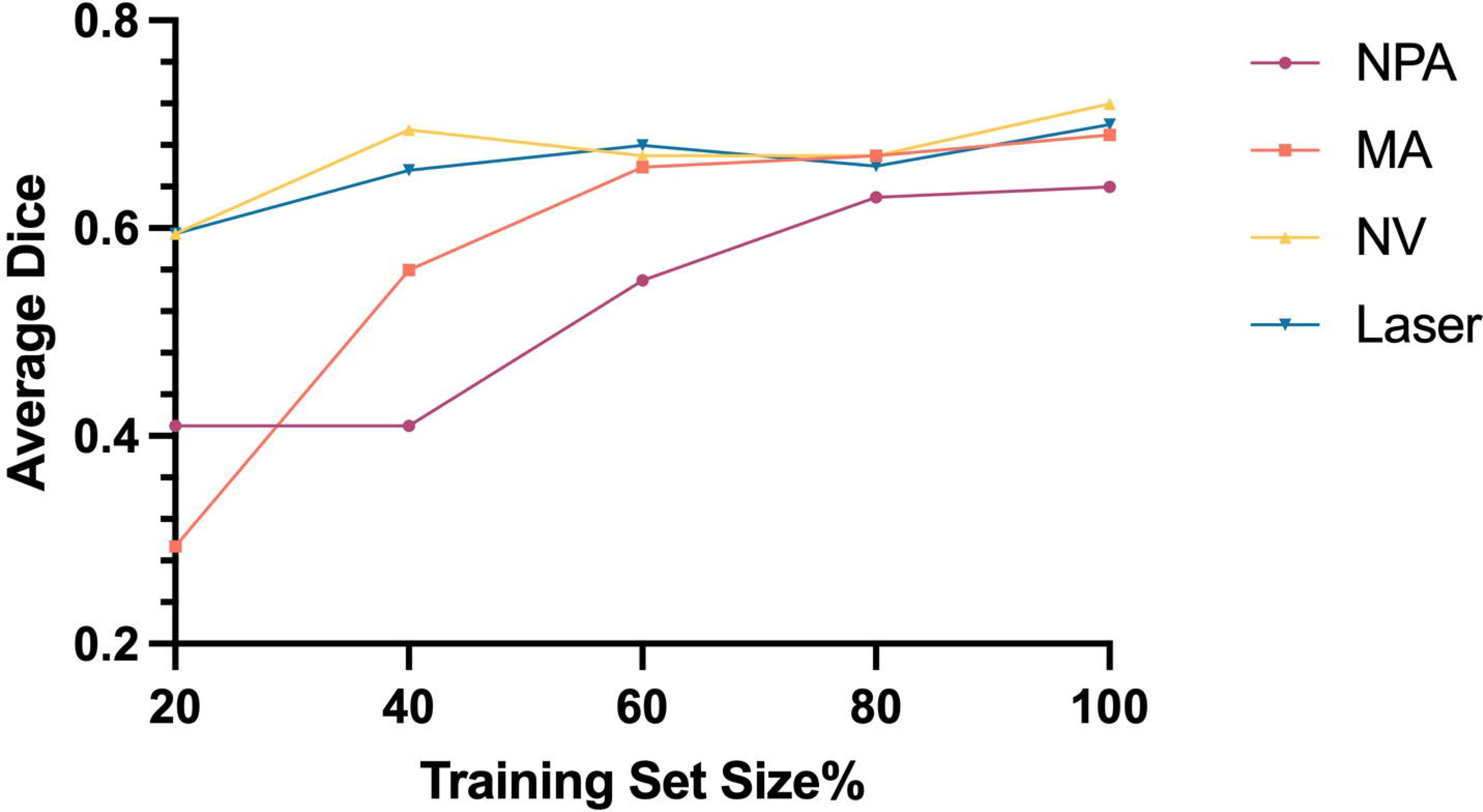
Influence of training set size on average Dice scores for lesion segmentation.

## Discussion

In this work, we developed a model to segment FFA lesions, capable of recognizing and segmenting a spectrum of pathologies, including NPA, MA, NV, and laser spots. This model not only matchs past segmentation performances for the 55° dataset but also ventures into the less-explored area of laser spot segmentation. Furthermore, our exploration into UWF-FFA images suggests the model’s ability to adapt learned features to UWF imaging, despite the challenges posed by the limited dataset size

Recent studies, summarized in Table 1, have focused on the segmentation of lesions in FFA images, as summarized previously in. Among these, Zhao et al reported the most notable NPA segmentation performance using a dataset of 750 BRVO and 545 DR images, achieving an F1 score of 0.90 for NPA segmentation, which decreased to 0.84 when including FFA images with laser treatment.^14^ Other studies primarily concentrate on single-pathology segmentation within a single modality (e.g. 30°, 55°, or UWF FFA) and single disease type (such as DR, RVO, wAMD, or CSC). These models often target NPA and less frequently consider secondary targets like NV in DR or leakage in CSC, rarely covering more than two disease types. Additionally, these studies often exclude images with moderate vitreous or pre-retinal hemorrhage, failing to reflect the complexity and variable quality of clinical FFA images, which affects model robustness. Our segmentation performance for NPA, MA, and NV are in line with those achieved in previous studies. For previously unexplored targets, the segmentation of laser treatment effects is satisfactory.

Despite the promising strides, our dataset’s limited UWF images, results in performance declines for NPA, MA, and NV compared to 55° images, though Laser performance remains consistent, suggesting possible learned features transferability. From a disease classification standpoint, the segmentation of NPA in RVO significantly outperformed that in DR, corroborating previous studies, possibly due to NPA’s more distinct continuity in BRVO.^15,18,19,21,23^ NV segmentation excelled in the venous and late phase over the arteriovenous phase, likely because some NV in our dataset just showcasing abnormal vascular networks and minimal or no leakage then. Our ablation study revealed that larger datasets correlate with higher Dice scores. Meanwhile, Zhao et al.’s study with the largest dataset (1295 images), achieved the highest NPA score, suggesting that larger datasets could enhance FFA segmentation performance.^14^ However, high-quality FFA annotations traditionally require extensive time and effort from ophthalmic specialists. Our approach could mitigate some of these challenges. Regarding annotator variability, Joan’s study reported a NPA Dice score of 0.65 between two annotators,^15^ while Lee reported IoUs of 0.793 and 0.625 for NP and NV labels respectively,^23^ highlighting the inevitable existence of intergrader variability. This level of variability closely matches the differences between our model’s predictions and the ground truth, suggesting that our model could potentially expand databases and alleviate the burden on graders.

On one hand, our model holds the promise of improving ophthalmic practice by offering automated analysis, aiding therapeutic decisions, and bolstering efficiency in clinical settings. It improves the interpretability of FFA examinations, crucial for ophthalmologists lacking specialized interpretive skills. Furthermore, measuring NPA is critical for predicting the diseases progression and guiding timely interventions. In CRVO, eyes with NPAs exceeding 10-disc areas on 55°FFA images are at a higher risk of developing anterior segment NV, ^27^ while an initial NPA over 75-disc areas on UWF FFA images greatly increases the risk of progressing to ischemic CRVO within a year.^28,29^ Identifying these high-risk patients early can improve their outcomes. Moreover, the model set the foundation for the interpretability of AI models, facilitating interpretable report generation and improving current ophthalmicquestion answering models.^30–33^ It may significantly contribute to the advancements in cross-modal image generation by offering high-quality, segmented lesion guidance, improving the generative models to produce realistic medical simulations for educational and diagnostic purposes.^34–36^

Our study has certain limitations. First, our dataset is restricted in size, with limited UWF images. Moreover, the absence of an external validation set may limit the generalizability of our findings. Second, the model currently utilizes FFA images only; adding paired color fundus images could improve accuracy, particularly in distinguishing NPA from HE—a valuable direction for future research. Lastly, our focus has been on a specific set of retinal vascular diseases, applying our model to other conditions may require further validation to confirm its broader clinical utility.

In conclusion, our study demonstrates the potential of a deep learning-based model to advance lesion segmentation in FFA images across a variety of retinal vascular diseases. The model exhibits overall satisfactory performance in segmenting multiple targets including NPA, MA, NV and laser spots. The model has the potential to expand databases, ease grader burden, standardize FFA image interpretation. Moreover, by enhancing interpretable AI, the model supports the development of sophisticated AI systems and contributes to cross-modal image generation. Future work may expand its utility and validation, promising to elevate ophthalmic care.

## Data Availability

All data produced in the present study are available upon reasonable request to the authors

---

**Figure S1.**
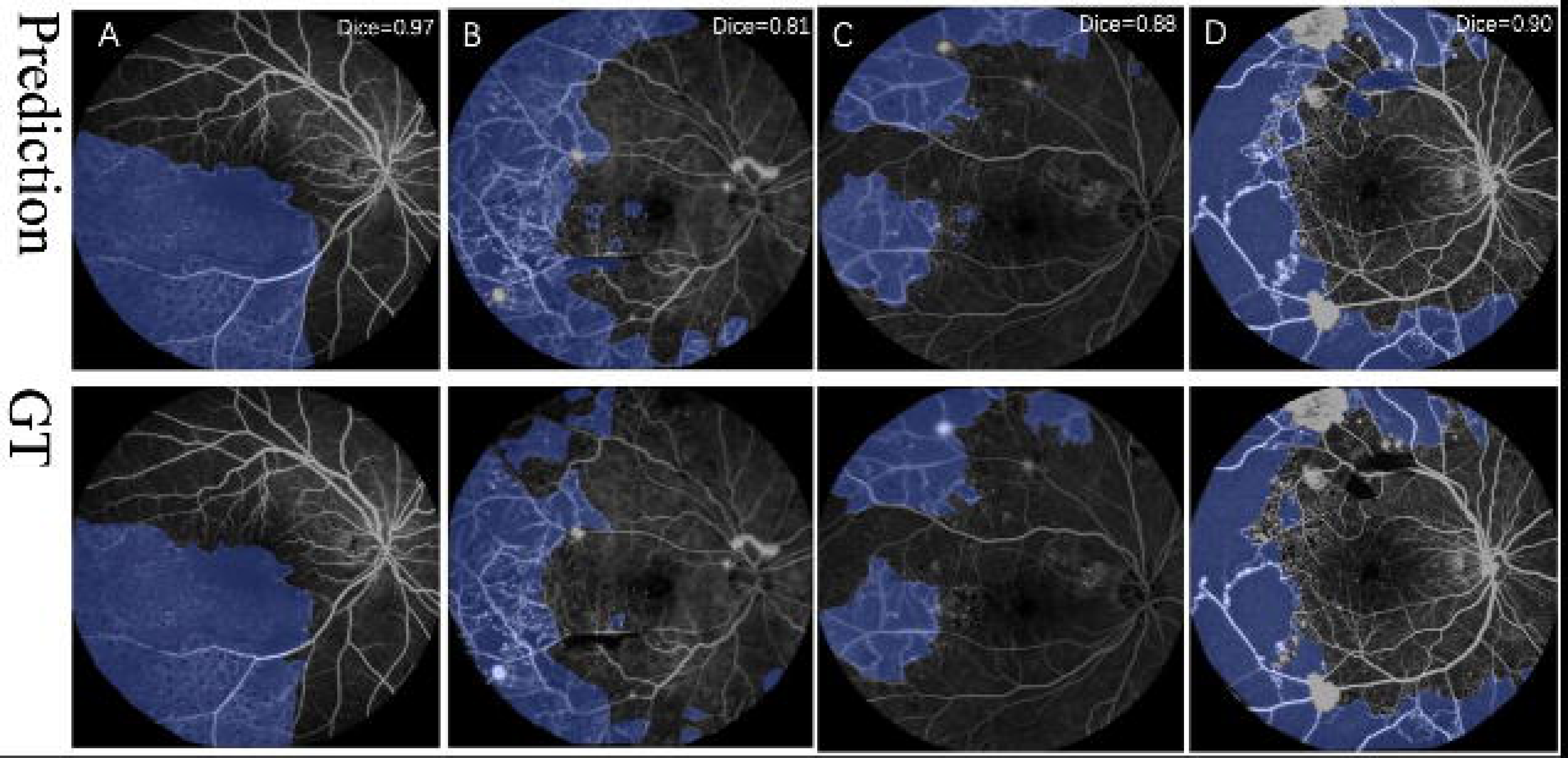
Visualization of NPA predictions versus ground truth in 55° FFA images.

**Figure S2.**
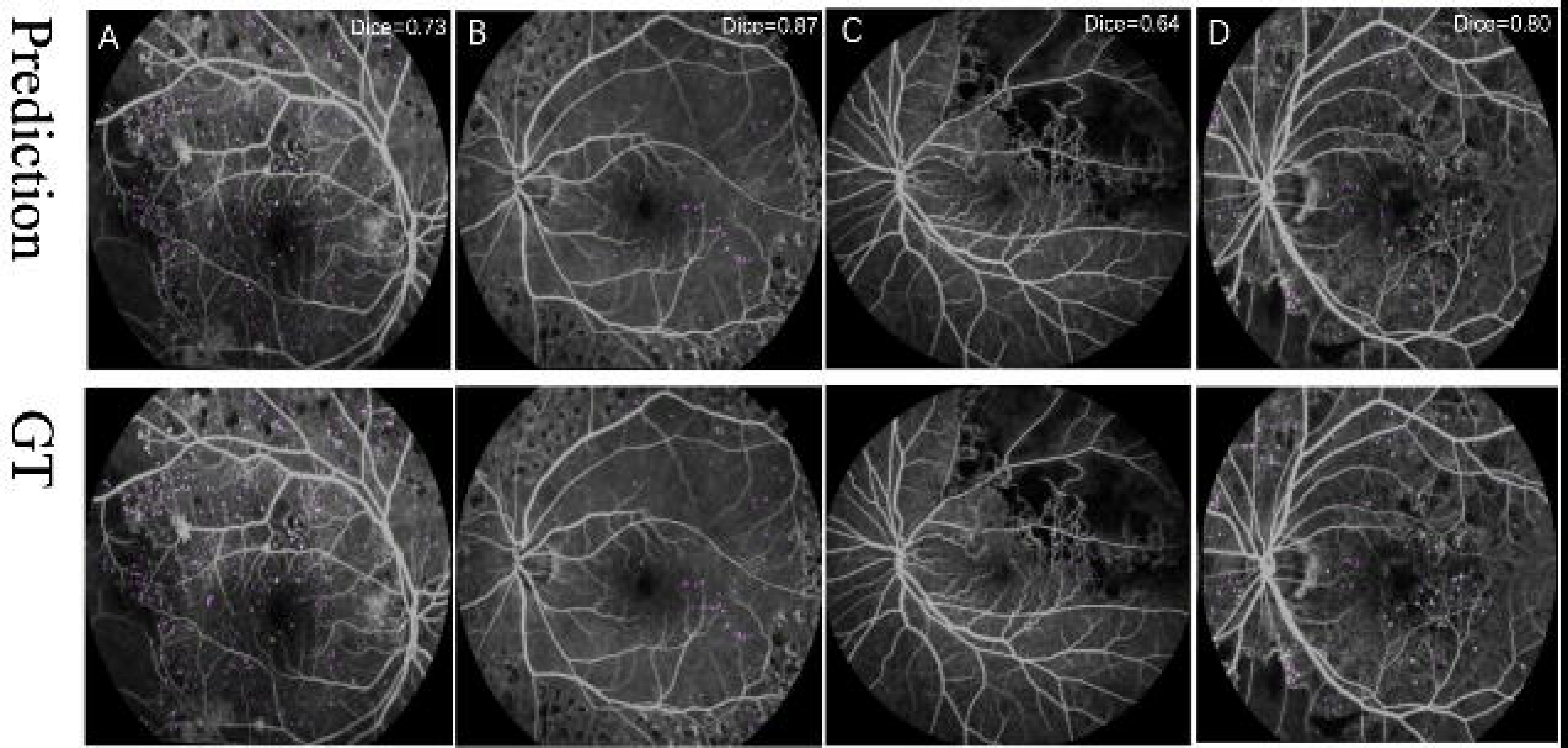
Visualization of MA predictions versus ground truth in 55° FFA images.

**Figure S3.**
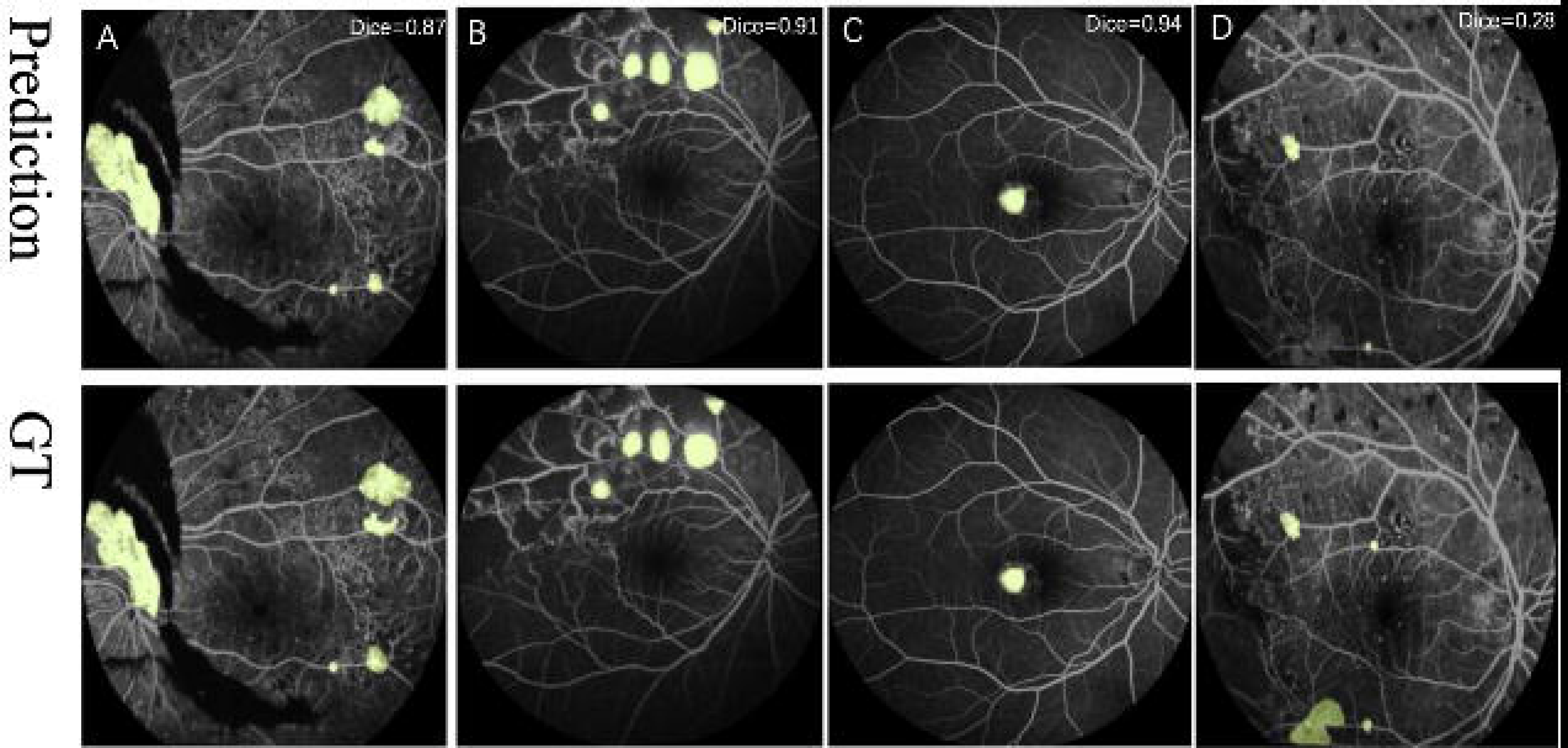
Visualization of NV predictions versus ground truth in 55° FFA images.

**Figure S4.**
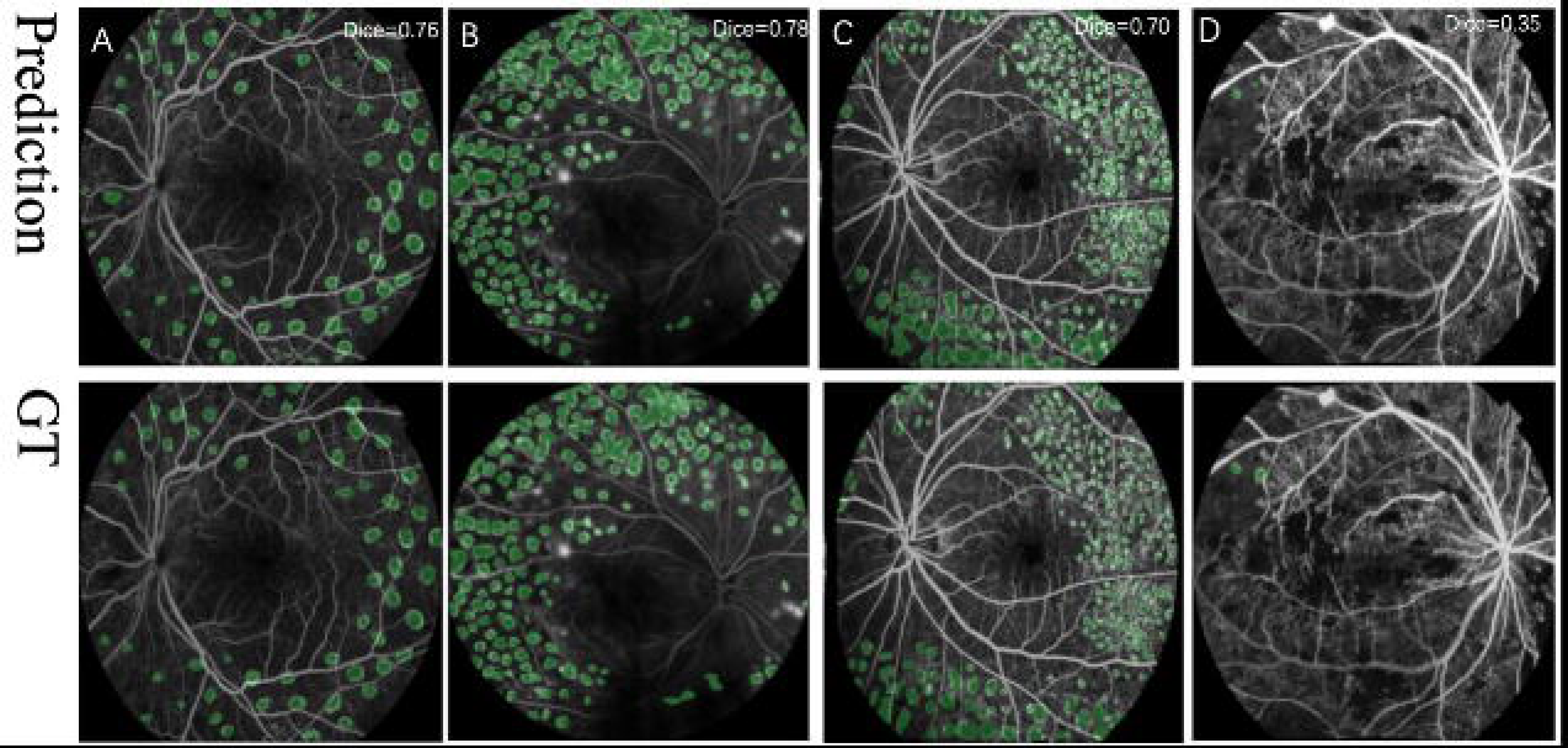
Visualization of laser spots predictions versus ground truth in 55° FFA images.

**Figure S5.**
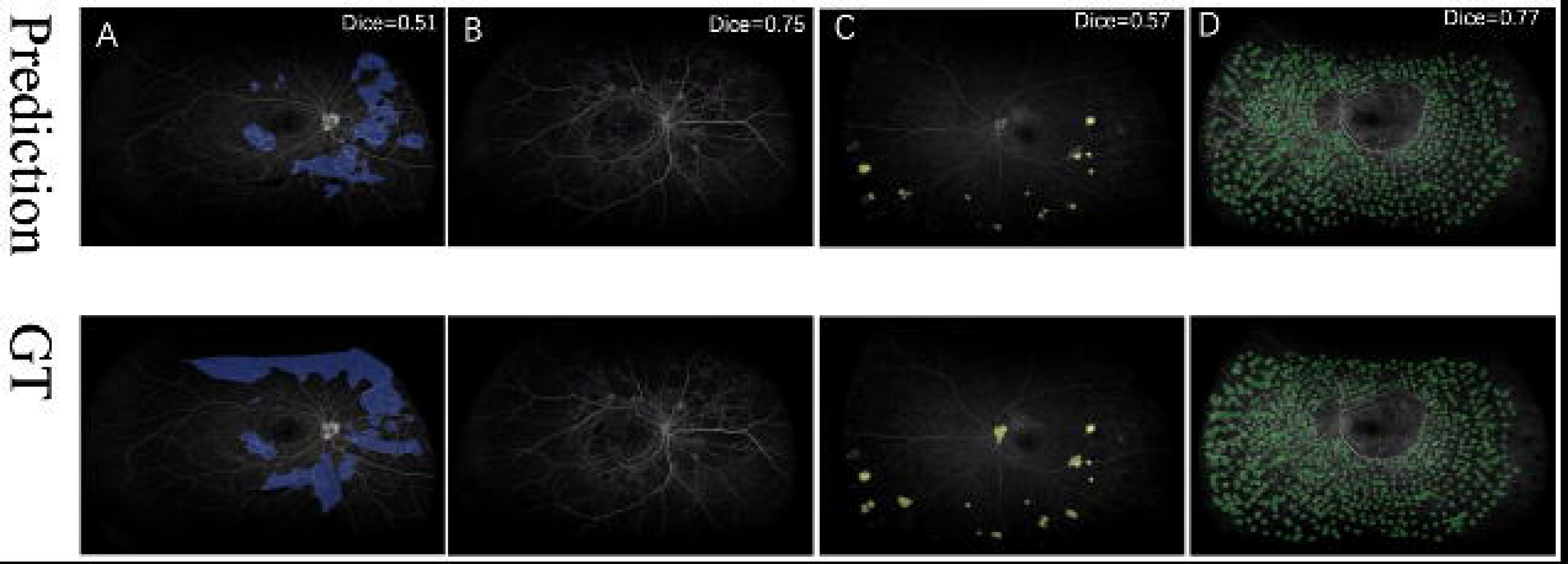
Visualization of target label predictions versus ground truth in UWF FFA images.

## Acknowledgments

We thank the InnoHK HKSAR Government for providing valuable supports.

Danli Shi and Mingguang He disclose support for the research and publication of this work from the Start-up Fund for RAPs under the Strategic Hiring Scheme from HKSAR (P0048623), the Global STEM Professorship Scheme (P0046113) and Henry G. Leong Endowed Professorship in Elderly Vision Health. The funders had no role in study design, data collection and analysis, decision to publish, or preparation of the manuscript.

## Commercial Relationship Disclosure

None

## Contributors

DS and MH conceived the study. SH and WZ built the deep learning model. ZZ, FS, and DS labeled or confirmed the annotations. ZZ did the literature search and analyzed the data. DS, ZZ, and FS contributed to key data interpretation. ZZ and SH wrote the manuscript. All authors critically revised the manuscript.

## Data sharing statement

The data used for model development of this study are not openly available due to reasons of privacy and are available from the corresponding author upon reasonable request.

